# Covid19 Pandemic Policy Monitor (COV-PPM): European level tracking of non-pharmaceutical interventions

**DOI:** 10.1101/2021.05.20.21257507

**Authors:** Kayvan Bozorgmehr, Sven Rohleder, Stella Duwendag, Amir Mohsenpour, Victoria Saint, Andreas Gold, Sukhvir Kaur, Niklas Nutsch, Diogo Costa

## Abstract

The Covid-19 Pandemic Policy Monitor (COV-PPM) prospectively documents the measures taken to contain SARS-Cov-2 transmission across countries in EU27, EEA and UK. In Germany, measures have also been documented at the federal state and, partially, at the district levels. Non-pharmaceutical interventions (NPIs) implemented since March 2020 have been retrieved and updated weekly from official governments webpages, Ministries of Health, National (Public) Health Institutes or Administrations. NPI categories refer to restrictions, closures or changes in functioning implemented in thirteen domains: public events (gatherings in indoor or outdoor spaces); public institutions (kindergartens, schools, universities); public transport (trains, buses, trams, metro); citizens movement/mobility (e.g. pedestrians, cars, ships); border closures (air, land or sea, all incoming travels, from high-risk regions, only non-nationals); measures to improve the healthcare system (e.g. human resources or technical reinforcement, redistribution, material or infrastructural); measures for risk/vulnerable groups (e.g. elderly, chronically ill, pregnant); economic measures (e.g. lay-off rules establishment, actions to avoid job-loss, tax relaxation); testing policies (e.g. testing criteria changes); nose and mouth protection rules, vaccination and others/miscellaneous measures.

## Background & Summary

The pandemic of Severe Acute Respiratory Syndrome Coronavirus Type 2 (SARS-Cov-2) demonstrated the lack of knowledge about the effectiveness of non-pharmaceutical interventions (NPIs) to mitigate or control transmissions at the population level. In the early stages of the pandemic, knowledge on NPI effectiveness was mostly limited and predominantly built on historical experience from previous influenza pandemic and SARS^1^. Recognizing such limitation, efforts were made by different institutions to monitor NPIs in different world regions.

In March 2020, we established the Covid-19 Pandemic Policy Monitor (COV-PPM) with the main goal of documenting the temporal sequence of the containment measures/NPIs being implemented in the EU27, EEA as well as in the UK at a national level, with higher resolution at federal state and (partially) district-level for Germany.

Other structured data collection efforts are being developed by different consortia, categorizing NPIs across the globe using different criteria and aggregation levels and updated with different periodicity. For example, the Response2covid^2^ considers 13 public health measures, and covers economic measures to a larger extent than the Oxford Tracker, and is updated monthly. The CoronaNet project^3^ covers 195 countries using a crowd-sourcing format, with volunteering applicants who answer a standardized questionnaire (survey design) about NPIs identified through different sources. They also partnered with a machine-learning company to automate their data collection through web-scrapping thousands of sources. The HIT-COVID tracker^4^ also covers 13 NPIs and resorts to volunteers who provide the data, focusing more extensively on the USA, but also with global coverage. Another NPIs tracker established by a consortia headed by a group from Vienna, Austria^5^ has been recording on a weekly basis measures implemented all over the world with four levels of sub-category disaggregation, thus making it a rich platform of information.

The European Observatory on Health Systems and Policies, has been developing a COVID-19 Health Systems Response Monitor (HSRM) that collects and organizes “one-off” information on how countries’ health systems are responding to the Pandemic crisis^6^. The HSRM is focused on measures taken by governments with a direct impact in health systems functioning, while other NPIs, such as border closures or general economic and social welfare protection measures for example, are recorded with less precision and categorized as “other measures”.

The University of Oxford has also been developing a COVID-19 Government Response Tracker (OxCGRT)^7^ to collect information on several common policy responses that governments (worldwide) have taken to respond to the pandemic, and compute a “Stringency Index” for comparative purposes, based on the restrictiveness of measures. These are all tools that might be useful for further testing the impact of NPIs implemented to tackle Covid-19 and evaluate the impact of measures considering other relevant outcomes.

COV-PPM shares some of the approaches and rationales of other trackers, but also differs from other efforts in its methodology, which aims to monitor NPIs prospectively, as they are being implemented, across 32 European nations, and with a special focus in Germany (where the research team is based), where NPIs are being documented at the federal state level and also to a limited extent in selected districts.

COV-PPM will allow answering multiple research questions related to the impacts of specific measures on the SARS-Cov-2 pandemic across different geographical regions or population subgroups and will also enable associations to be tested between higher-level characteristics (e.g., macro-economic indicators) and NPI effectiveness, accounting for other contextually relevant characteristics of the population.

In this work, we describe the design, methods and procedures taken for the establishment of the COV-PPM, and we present descriptive results of NPIs recorded until December 2020 in the EU-27, EEA, the UK and in Germany’s 16 federal states.

## Methods

Starting in March 2020, the Covid-19 Pandemic Policy Monitor (COV-PPM) was established to record on a weekly basis, the NPIs taken in the EU27, EEA and UK (total of 32 countries). Additionally, data for Germany are being collected also at the federal state level (Nomenclature des Unités Territoriales Statistiques, NUTS2) and partially at the district-level (NUTS3). A team of researchers and research assistants at Bielefeld University in collaboration with scientists from Heidelberg University Hospital, collected and coded the NPIs being implemented at the national level in these countries, and at federal- and district-levels in Germany, using a pre-established Microsoft Excel form for data entry.

Official national health authorities’ websites are considered primary sources (e.g. Ministries of Health, National Public Health Institutes and any specific websites managed by national governments to convey information about Covid-19), followed by national news websites. For German federal states and districts, information from health authorities of the corresponding geographical level is included as well. The full list of sources is provided in Panel 1. Google translate was used for website translation whenever English versions of websites were not available (and the team of researchers/assistants included German, French, Spanish and Portuguese speakers, that helped understand and document specific elements whenever translations provided dubious wording).

### Panel 1

#### List of sources used to document non-pharmaceutical interventions being implemented in COV-PPM

##### Countries

Austria: https://www.sozialministerium.at/Informationen-zum-Coronavirus/Coronavirus---Haeufig-gestellte-Fragen.html

Belgium: https://www.info-coronavirus.be/en/ https://www.vlaanderen.be/gezondheid-en-welzijn/gezondheid/gezondheid-en-preventie-tijdens-de-coronacrisis https://coronavirus.brussels/index.php/en/ http://www.ejustice.just.fgov.be/cgi/welcome.pl https://www.wallonie.be/fr/actualites/coronavirus-covid-19-mesures-regionales

Bulgaria: https://www.mh.government.bg/bg/novini/aktualno/

Croatia: https://www.koronavirus.hr/ https://www.koronavirus.hr/najnovije/34

Cyprus: https://www.pio.gov.cy/coronavirus/

Czech Republic: https://koronavirus.mzcr.cz/ https://www.vlada.cz/scripts/detail.php?pgid=103&conn=3151&pg=1 https://www.vlada.cz/cz/epidemie-koronaviru/dulezite-informace/mimoradna-opatreni-_-co-aktualne-plati-180234/

Denmark: https://um.dk/en/ https://coronasmitte.dk/en https://www.sst.dk/en/English https://www.ssi.dk/aktuelt/sygdomsudbrud/coronavirus https://en.ssi.dk/ https://coronasmitte.dk/en/overview

Estonia: https://www.sm.ee/et https://www.kriis.ee/et https://www.valitsus.ee/et/eriolukord-eestis#meetmed https://kkk.kriis.ee/et/kkk/fookuses-koroonakriisist-jarkjargult-valjatulek

Finland: https://thl.fi/en/web/infectious-diseases/what-s-new/coronavirus-covid-19-latest-updates https://valtioneuvosto.fi/en/information-on-coronavirus https://stm.fi/

France: https://www.gouvernement.fr/info-coronavirus https://solidarites-sante.gouv.fr/ https://www.santepubliquefrance.fr/

Germany: https://www.bundesregierung.de/breg-de/themen/coronavirus https://www.bundesgesundheitsministerium.de/service/gesetze-und-verordnungen.html

Greece: https://www.moh.gov.gr/articles/ministry/grafeio-typoy/press-releases https://www.moh.gov.gr/articles/health/dieythynsh-dhmosias-ygieinhs/metra-prolhpshs-enanti-koronoioy-sars-cov-2/ https://eody.gov.gr/category/epikairotita/ https://www.civilprotection.gr/el https://covid19.gov.gr/category/proliptika-metra-gia-tin-pandimia/ https://www.civilprotection.gr/el/announcements https://covid19.gov.gr/

Hungary: http://abouthungary.hu/blog/pm-orban-announced-extension-of-economy-protection-action-plan/

Iceland: https://www.government.is/news/ https://www.landlaeknir.is/koronaveira/thad-sem-thu-tharft-ad-vita/ https://www.covid.is/

Ireland: https://www.gov.ie/en/news/

Italy: http://www.salute.gov.it/portale/nuovocoronavirus/ https://www.gazzettaufficiale.it/dettaglioArea/12 http://www.salute.gov.it/portale/news/p3_2.html

Latvia: https://www.mk.gov.lv/lv/covid-19 https://covid19.gov.lv/index.php/en https://www.vm.gov.lv/lv

Liechtenstein: https://www.regierung.li/coronavirus https://www.llv.li/medienmitteilungen https://hebensorg.li/ https://tourismus.li/unser-land/ueber-liechtenstein/aktuelle-informationen-zu-coronacovid-19-im-fuerstentum-liechtenstein/ https://www.wko.at/service/aussenwirtschaft/coronavirus-newsticker-schweiz.html https://www.liechtenstein.li/land-und-leute/gesellschaft/gesundheitswesen/corona-virus/

Lithuania: https://lrv.lt/en/news https://koronastop.lrv.lt/en/ https://urm.lt/default/en/important-covid19

Luxembourg: https://covid19.public.lu/de/communications-officielles.html https://gouvernement.lu/fr/actualites/toutes_actualites.html https://data.public.lu/en/datasets/covid-19-rapports-hebdomadaires/

Malta: https://deputyprimeminister.gov.mt/en/Pages/health.aspx

Netherlands: https://www.rijksoverheid.nl/onderwerpen/coronavirus-covid-19/nieuws

Norway: https://www.regjeringen.no/en/whatsnew/finn-aktuelt2/id2415244/?topic=2692388 https://www.regjeringen.no/en/topics/koronavirus-covid-19/timeline-for-news-from-norwegian-ministries-about-the-coronavirus-disease-covid-19/id2692402/ https://www.fhi.no/en/ http://www.norwaynews.com/

Poland: https://www.gov.pl/web/coronavirus https://www.gov.pl/web/koronawirus/dzialania-rzadu https://www.gov.pl/web/coronavirus/travel https://www.gov.pl/web/coronavirus/temporary-limitations

Portugal: https://dre.pt/legislacao-covid-19-upo https://covid19estamoson.gov.pt/medidas-excecionais/

Romania: https://stirioficiale.ro/

Slovakia: https://www.korona.gov.sk/covid-19-usmernenia-obcania.php https://www.standardnepostupy.sk/klinicky-protokol-spdtp-klinicky-manazment-podozrivych-a-potvrdenych-pripadov-covid-19/

Slovenia: https://www.gov.si/en/news?date=&nrOfItems=20&start=0&tag%5B0%5D=554

Spain: https://administracion.gob.es/ https://www.lamoncloa.gob.es/lang/en/Paginas/index.aspx

Sweden: https://www.folkhalsomyndigheten.se/smittskydd-beredskap/utbrott/aktuella-utbrott/covid-19/ https://www.government.se/government-policy/the-governments-work-in-response-to-the-virus-responsible-for-covid-19/

Switzerland: https://www.bag.admin.ch/bag/en/home.html https://www.admin.ch/gov/en/start/documentation/media-releases.html?dyn_startDate=01.01.2015&dyn_organization=1

UK: https://www.gov.uk/search/news-and-communications https://www.england.nhs.uk/coronavirus/ https://www.wikiwand.com/en/COVID-19_pandemic_in_the_United_Kingdom

##### German federal states

Baden-Württemberg: https://www.baden-wuerttemberg.de/de/service/aktuelle-infos-zu-corona/aktuelle-corona-verordnung-des-landes-baden-wuerttemberg/

Bavaria: https://www.stmgp.bayern.de/coronavirus/rechtsgrundlagen/

Berlin: https://www.berlin.de/corona/massnahmen/verordnung/

Brandenburg: https://www.landesrecht.brandenburg.de/dislservice/public/index.jsp; https://brandenburg-impft.de/bb-impft/de/aktuelles/; https://mik.brandenburg.de/mik/de/start/service/presse/pressemitteilungen/

Bremen: https://www.gesetzblatt.bremen.de; https://www.amtliche-bekanntmachungen.bremen.de

Hamburg: https://www.luewu.de/gvbl/; https://www.hamburg.de/allgemeinverfuegungen/

Hesse: https://www.hessen.de/fuer-buerger/corona-hessen/verordnungen-und-allgemeinverfuegungen; https://www.hessen.de/fuer-buerger/corona-in-hessen/interviews-reden-und-mehr/corona-massnahmen-der-landesregierung-seit-dezember-2020

Lower Saxony: https://www.niedersachsen.de/Coronavirus/vorschriften/vorschriften-der-landesregierung-185856.html; https://www.niedersachsen.de/Coronavirus; https://www.niedersachsen.de/Coronavirus/aktuelle-presseinformationen-186247.html#aeltere_Meldungen

Mecklenburg-Vorpommern: https://www.regierung-mv.de/corona/Verordnungen-und-Dokumente/; https://www.regierung-mv.de/corona/#wichtige%20Dokumente; https://www.regierung-mv.de/Aktuell; https://www.lagus.mv-regierung.de/Gesundheit/InfektionsschutzPraevention/Impfen-Corona-Pandemie/

North-Rhine-Westphalia: https://www.mags.nrw/coronavirus-rechtlicheregelungen-nrw; https://www.land.nrw/nl/node/23375

Rhineland-Palatinate: https://corona.rlp.de/de/startseite/; https://corona.rlp.de/de/service/rechtsgrundlagen/

Saarland: https://www.saarland.de/DE/portale/corona/service/rechtsverordnung-massnahmen/rechtsverordnung-massnahmen_node.html

Saxony: https://www.coronavirus.sachsen.de/amtliche-bekanntmachungen.html; https://www.coronavirus.sachsen.de/index.html; https://www.coronavirus.sachsen.de/newsroom-4155.html

Saxony-Anhalt: https://ms.sachsen-anhalt.de/themen/gesundheit/aktuell/coronavirus/

Schleswig-Holstein: https://www.schleswig-holstein.de/DE/Landesregierung/Themen/GesundheitVerbraucherschutz/Coronavirus/coronavirus.html; https://www.schleswig-holstein.de/DE/Schwerpunkte/Coronavirus/_documents/teaser_erlasse.html;jsessionid=F274B822369590071BEF2060DEBF64AE.delivery1-replication; https://www.schleswig-holstein.de/DE/Schwerpunkte/Coronavirus/Presse/PI/pressemitteilungen_node.html

Thuringia: https://www.tmasgff.de/covid-19/rechtsgrundlage https://corona.thueringen.de/

The categories of NPIs scoped were developed based on the knowledge of current measures being implemented in different countries and contexts and complemented with the policies detailed in previous WHO reports and guidance (which are mainly based on the policies taken during the 2009 H1N1 influenza pandemic)^1^. The categories were progressively refined and adapted as the Covid-19 pandemic unfolded and different measures appeared (e.g. masks or mouth and nose protection, and vaccination rules and procedures). For each NPI category the team coded the exact dates of implementation and provided a textual description of the relevant information needed to contextualize each measure, saving the respective source (saved to pdf format, ensuring traceability of the collected information, since official health authorities websites, for example, are updated regularly). The categories established comprehensively cover several societal domains and regional resolutions, as detailed in Table 1.

**Table 1.**
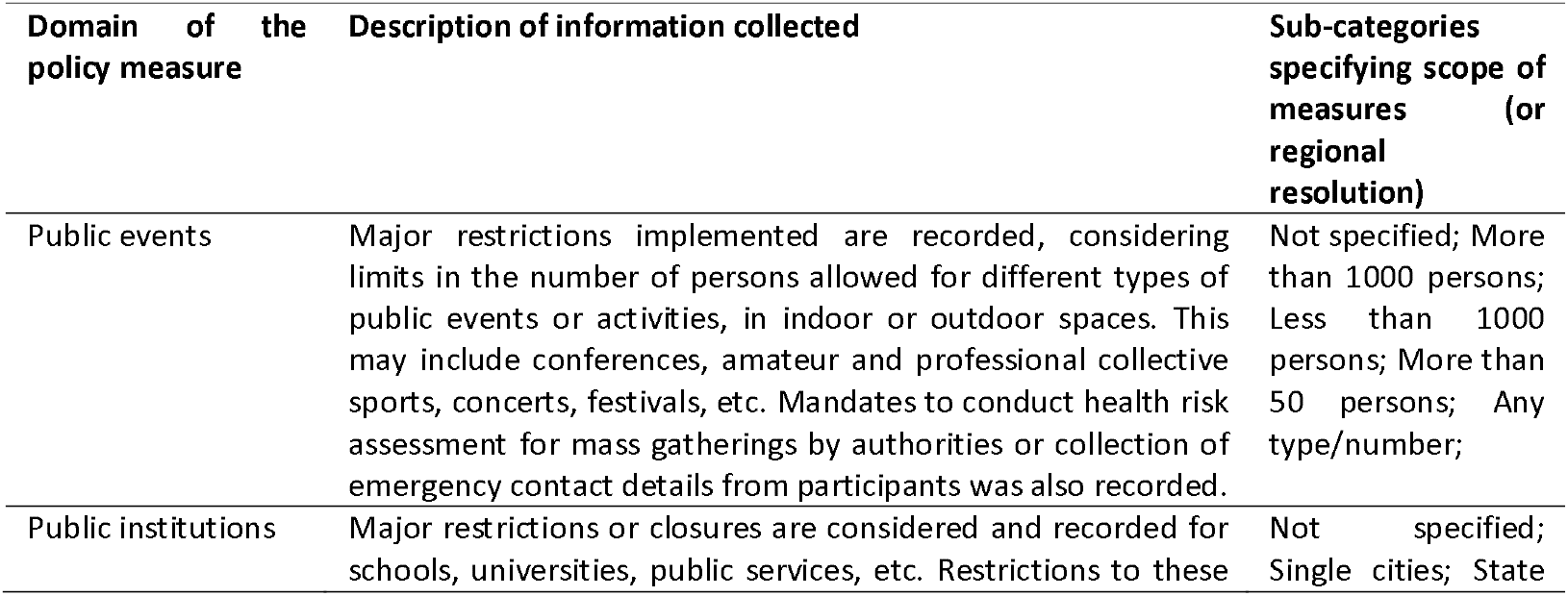
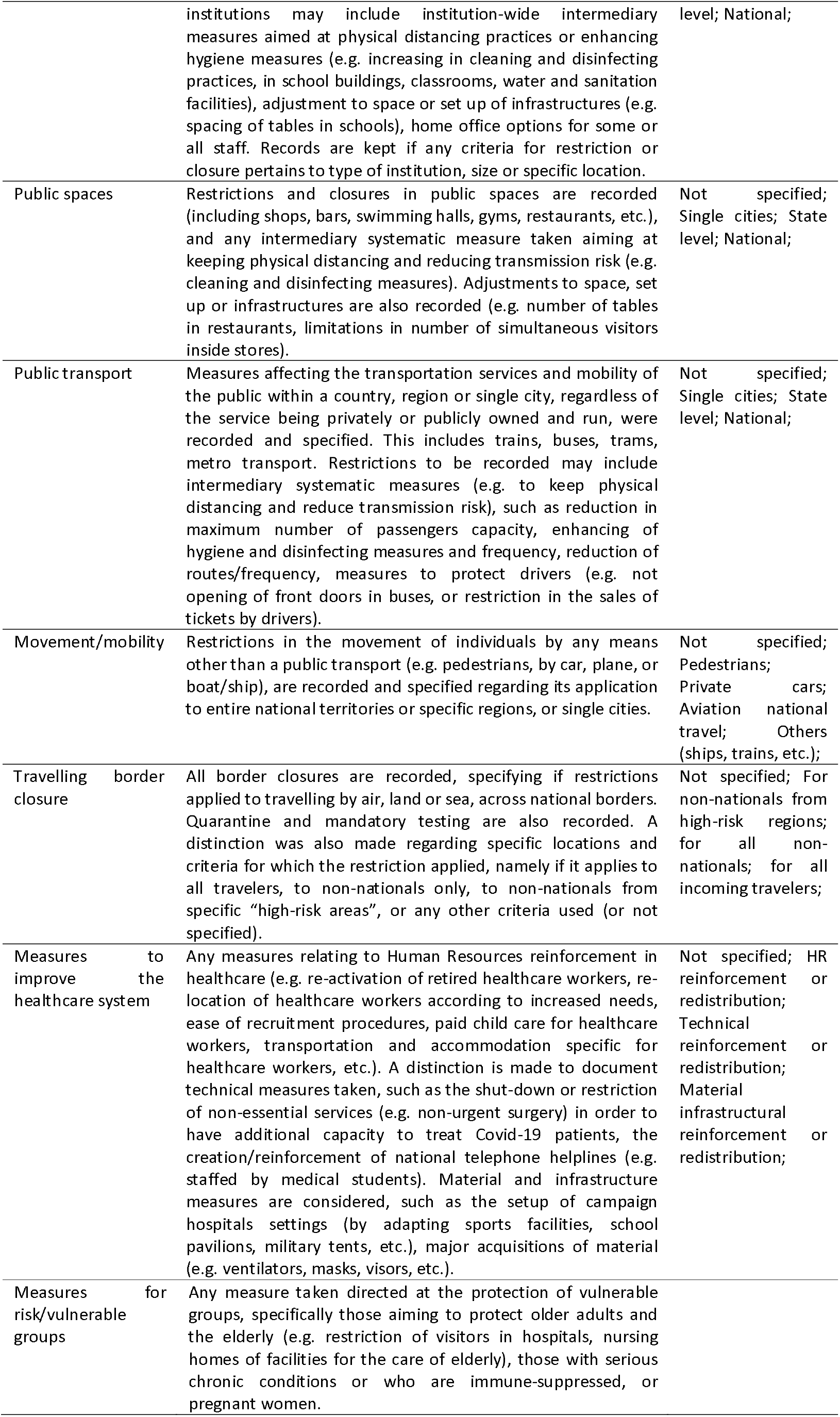
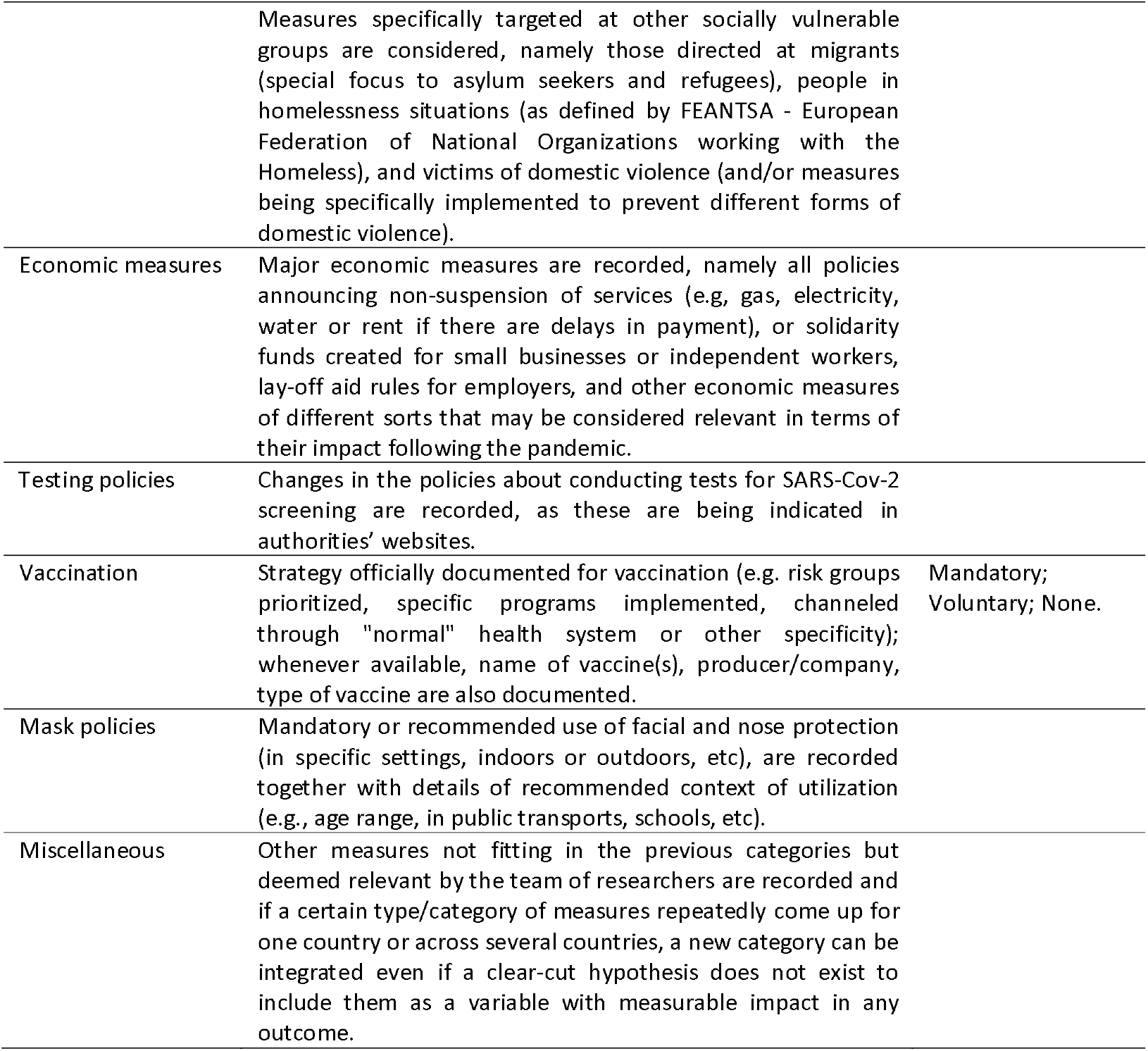
Non-pharmaceutical interventions monitored by the COV-PPM in EU-27, EEA, UK and in the German Federal States.

The textual description added to each measure at the time of their coding, was preceded by selected key-words to allow qualification and recoding of NPIs according to their increasing or decreasing stringency. Specifically, for each point in time attached to the start, change or stop of each NPI category, a qualifier was added to identify and count if the coding refers to an additional “restriction”, “withdrawal” or “recommendation”. For the specific case of Masks utilization, two identifiers were added also to the categories where mouth and nose protection were additionally implemented as mandatory or recommended (e.g. enforcement of masks utilization in public transports, public institutions, public events).

## Data Records

The latest version of data (updated until 14 December 2020) are available on the Harvard Dataverse Repository (https://doi.org/10.7910/DVN/ANTOH7). Data visualizations and a description of the project are available online at https://www.uni-bielefeld.de/fakultaeten/gesundheitswissenschaften/ag/ag2/covid19.xml. The database is available in long format, with each column representing the NPI domains and each code representing a degree of implementation or regional scope of application. The COV-PPM codebook is detailed in Table 2.

**Table 2.**
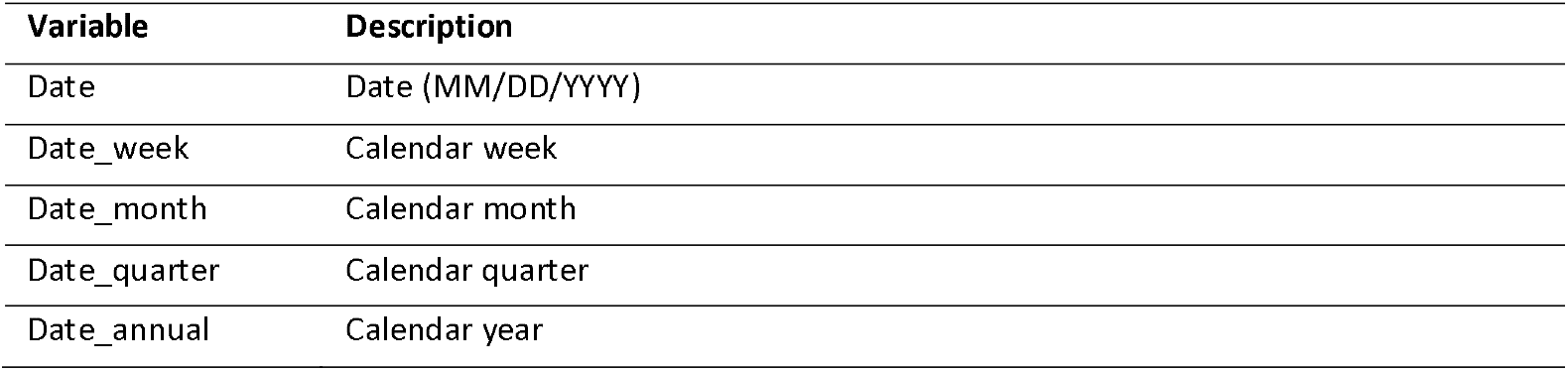
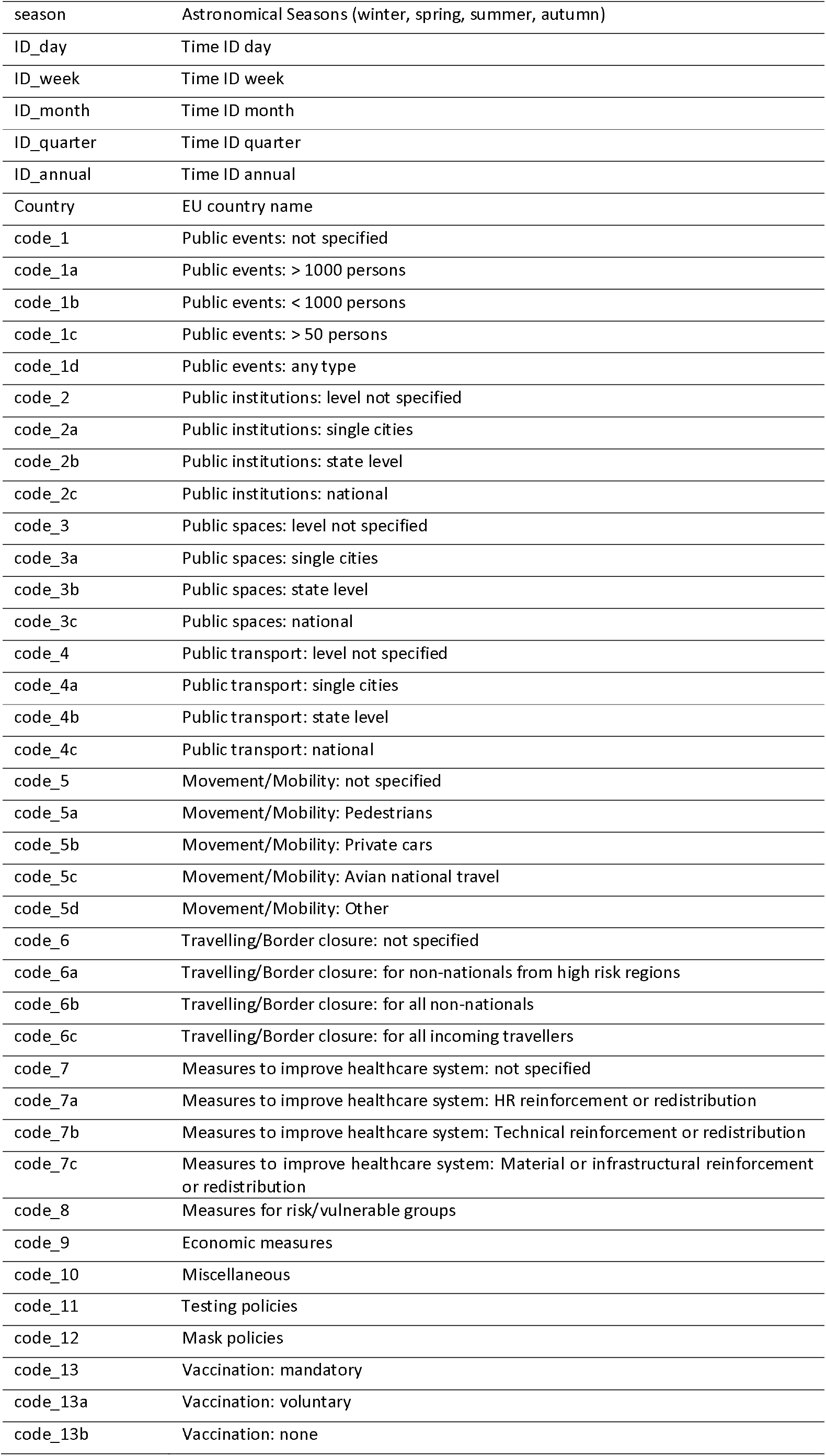
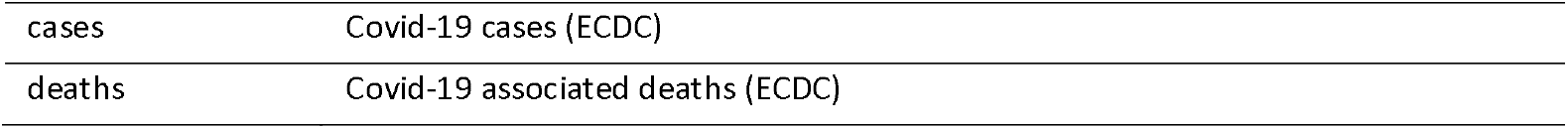
Variable description in COV-PPM

### Elements added in the dataset for Germany

In addition, the analysis of the textual elements and qualifiers (i.e., key-words identifying “restriction” or “withdrawal”) retrieved with each NPI description for the 16 German federal states, allowed the creation of a subset of variables with disaggregation of broader selected categories: for public institutions, distinct variables identifying measures specifically aiming at a) schools, b) kindergartens and c) universities/higher education, d) gastronomy and e) nightlife facilities. The applied variable coding to restrictions in public institutions is detailed in Appendix. The same additions are being developed for application at the EU-data level.

## Technical Validation

The COV-PPM was set-up to include detailed information regarding specific aspects of the categories of measures being monitored, by adding a textual description for each measure registered and qualifying with selected “buzzwords” each change in the NPIs, prospectively. The team of researchers and assistants collecting the data, conducted frequent consensus meetings (weekly) and continuously updated the protocol for data collection according to changes also occurring throughout Europe, to encompass all nuances of increased or decreased stringency being implemented.

A Quality Assessment (QA) exercise was also conducted for the data gathered at the EU-level, and at the federal and district levels in Germany. The QA procedure consisted of a quantitative component and a qualitative one.

In the quantitative component, the team plotted the measures collected using interactive graphs, allowing a simple visualization of the data (example provided in Figure 1). Based on these plots, the team search for individual data points for each NPI to check for their accuracy and for potential non-plausible temporal sequences of NPIs (e.g. restrictions to public events or public transports coded for very short periods or with several interruptions in the time-series, measures coded before February 2020).

**Figure 1.**
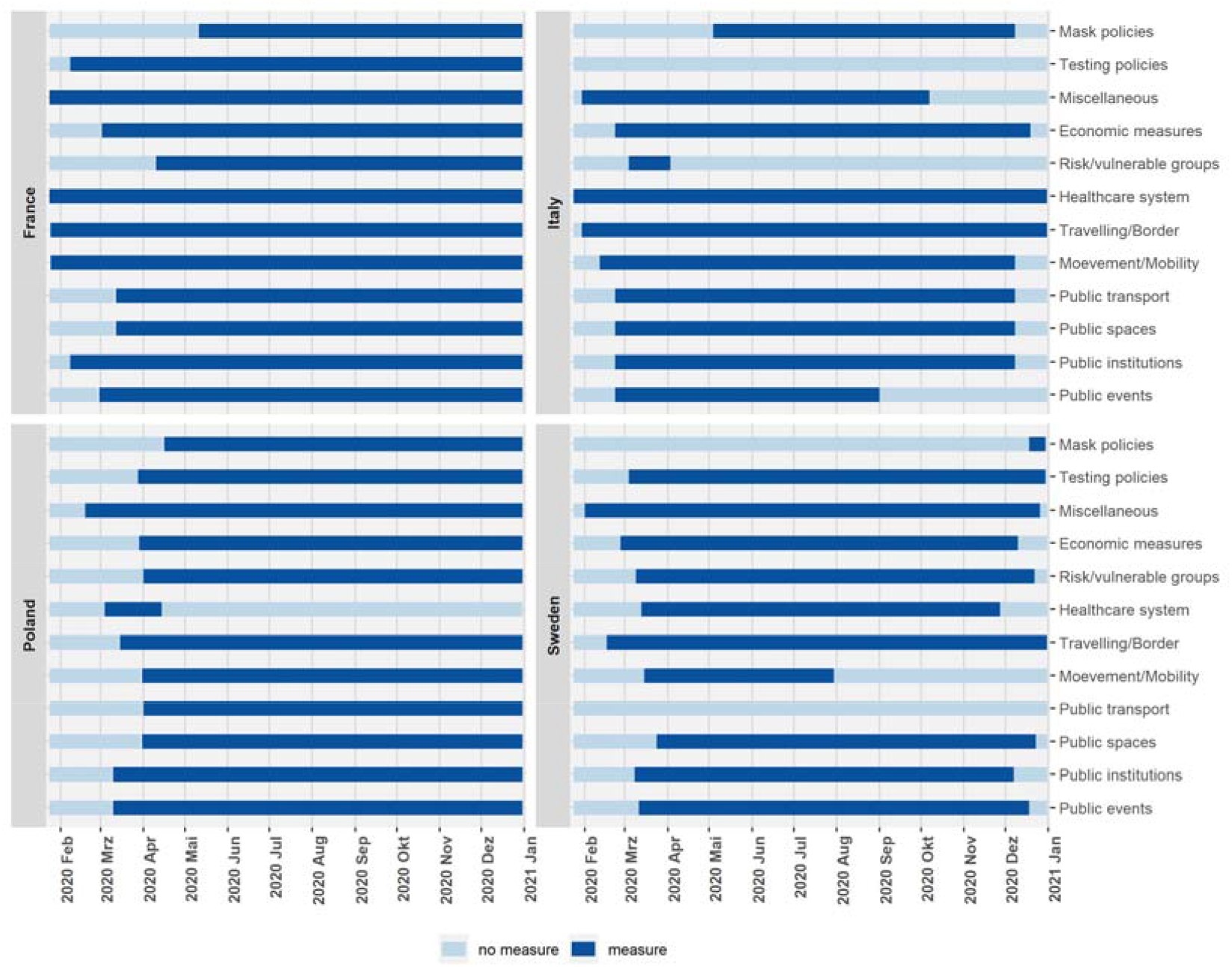
Example of bar graph used to describe data gathered to guide the Quality Assessment of COV-PPM.

Following identification of potential coding mistakes visualized in the graphs, the individual country or federal state datasets were checked for the coding and a textual description recorded to assess whether an error was made in the documentation.

The qualitative component of the QA procedure consisted in analysing the numerical coding and comments/textual elements saved for about 5-10% (around 2 weeks) of the total time sequence of each NPI, for each country, German federal state, and districts. The selection of the proportion of time to examine was based on the amount of comments/textual information registered (sequences with more frequent elements were selected but could vary for each NPI and for each geographical unit). The numerical coding and textual information were then compared with the saved source to identify mismatches between the information provided in the sources and in the database. Two independent reviewers completed the QA process described for the German federal states, while most of the QA procedure at the EU level was conducted by a single reviewer. Incongruencies found were then corrected.

At time of writing this text, a qualitative assessment of the text included for each subcategory is also being conducted, in order to establish even more fine-graded categories of measures that emerge across the different countries and German federal states, thus adding a layer of specificity to the future analysis potential. Analysis of this qualitative information gathered, which is retrieved from countries’ official governmental sources, allows disentangling the change that occurred to the nature of some NPIs, from complete closures to conditions under which specific public or private spaces can operate or host events, for example (which would have been aggregated as the same measure, otherwise). This also provides the needed flexibility to compare the data gathered with other existing public policy trackers for Covid-19, allowing for cross-validation.

Such a cross-validation exercise is also being implemented at the German federal state level by comparing the data gathered with the one provided by the National Ministry for Economy and Energy (www.corona-datenplattform.de/) and the results can be shared upon reasonable request.

## Data Availability

The dataset along with codes (prepared with R) to generate analytical variables can be accessed via Harvard Dataverse repository (https://doi.org/10.7910/DVN/ANTOH7). The data and code is for public, scientific, non-commercial use and can be retrieved upon reasonable request by submitting a proposal to.

## Usage Notes

The data gathered in the scope of the COV-PPM can be used in different ways to describe the NPIs duration and combination across the EU-27, the UK and EEA and at the German federal state level. Examples of such utilizations can be:

a. As categorical data with horizontal bars over time by country (Figure 2);
b. As percentage of observation time in which individual NPIs are in place in each country (or German federal state);
c. As categorical data descriptively overlayed with incidence data in each country (or German federal state);
d. As an index using combinations of NPIs simultaneously implemented at different sub-categories or levels, i.e., deriving new variables for each NPI, coded as 1, 2, 3, 4 (or 5), summarizing the simultaneous existence of the sub-categories “not-specified”, “a”, “b”, “c” or “d” (or “e”) respectively, for each of the NPIs described in Table 1. Example for the NPI reflecting restrictions in “Public Events” would be, code “a = more than 1000 persons”, code “b = less than 1000 persons”, code “c = more than 50 persons”, code “d = any type/number”.

## Code Availability

**Figure 2.**
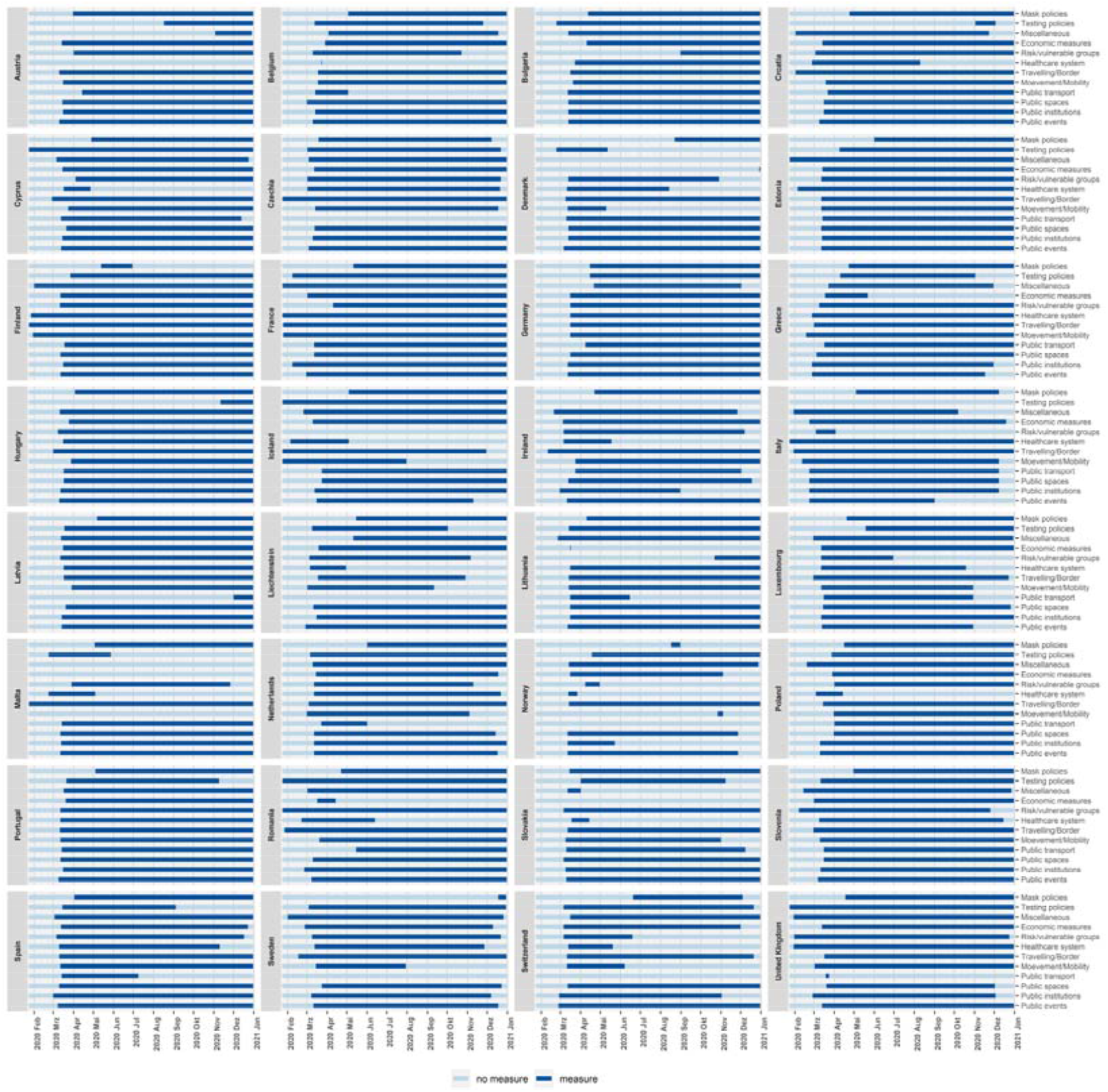
Example of horizontal display of categorical data gathered.

## Acknowledgements

We acknowledge all colleagues involved in data collection from the beginning of this project: Judith Wenner, Rosa Jahn, Anna Mundry, Cathrin Stöber, Helena Haskia, Johanna Nonte, Catharina Mühling, Leonie Klose, Lilian Salis, Sarah Markgraf, Neelab Hakim, Julia Winkelnkemper, Fatemeh Ebrahimi, Nicole Krzisczyk, Johanna Landes, Annalena Sander, Oxana Klassen, Valentina Klapp. We further acknowledge conceptual contributions from Dr Matthias an der Heiden and Dr Viviane Bremer from the Robert Koch Institute, regarding approaches to refine the coding.

We acknowledge financial support by the Rectorate of Bielefeld University, and the Federal Ministry of Health Germany (ZMV I 1 - 25 20 COR 410).

## Author contributions

KB: Conceived the CoV-PPM; Study design; Methods; Writing; General Oversight; Funding acquisition

SR: Study design; Methods; Writing; Data Management; Data Collection; Project Coordination

SD: Study design; Methods; Writing; Data Management; Data Collection; Project Coordination

AM: Study design; Methods; Writing; Data Collection

VS: Study design; Methods; Writing; Data Collection

AG: Study design; Methods; Writing; Data Collection

SK: Study design; Methods; Writing; Data Collection

NN: Study design; Methods; Writing; Data Collection

DC: Study design; Methods; Writing; Data Management; Data Collection; Project Coordination; General oversight.

## Appendix

**Table A1.** Codebook with the variables created to disaggregate information about major restrictions to the functioning of public institutions identified in the scope of COV-PPM.

| Category | Item | Variable | Variable value | Description |
| --- | --- | --- | --- | --- |
| Public institutions | School | school | 0 = no measure<br>1 = restricted measure<br>2 = relaxed measure<br>3 = both measures in place | General restricted and relaxed measures in schools.<br>"3 = both measures in place" means that both restricted and relaxed measures are present in schools at the same time. |
|  |  | school_prim | 0 = no measure<br>1 = restricted measure<br>2 = relaxed measure<br>3 = both measures in place | General restricted and relaxed measures in primary schools.<br>"3 = both measures in place" means that both restricted and relaxed measures are present at the same time. |
|  |  | school_sec | 0 = no measure<br>1 = restricted measure<br>2 = relaxed measure<br>3 = both measures in place | General restricted and relaxed measures in secondary schools.<br>"3 = both measures in place" means that both restricted and relaxed measures are present at the same time. |
|  |  | school_res | 0 = no measure<br>1 + n = cumulative number of restricted measures | Cumulative number of restricted school measures over time.<br>The first restriction is given the value 1. All subsequent new restrictions become 1 + n number of previous restrictions. |
|  |  | school_mit | 0 = no measure<br>1 + n = cumulative number of relaxed measures | Cumulative number of relaxed school measures over time.<br>The first relaxation is given the value 1. All subsequent new relaxations come 1 + n number of previous relaxations. |
|  |  | school_res_mit | 1 = zero point | Interval scaled variable measuring the change in present restricted and relaxed measures. If there is a restriction, the value is increased by 0.01 points or decreased by this value if there is a relaxation. |
|  |  | school_closure | 0 = no measure in place<br>1 = closure<br>2 = opening | Categorical measure of school closures. |
|  |  | school_prim_closure | 0 = no measure in place<br>1 = closure<br>2 = opening | Categorical measure of primary school closures. |
|  |  | school_sec_closure | 0 = no measure in place<br>1 = closure<br>2 = opening | Categorical measure of secondary school closures. |
| Public institutions | **Kindergarten** | kita | 0 = no measure<br>1 = restricted measure<br>2 = relaxed measure<br>3 = both measures in place | General restricted and relaxed measures in kindergarten and nursery schools.<br>"3 = both measures in place" means that both restricted and relaxed measures are present at the same time. |
|  |  | kita_res | 0 = no measure<br>1 + n = cumulative number of restricted measures | Cumulative number of restricted kindergarten and nursery school measures over time. The first restriction is given the value 1. All subsequent new restrictions become 1 + n number of previous restrictions. |
|  |  | kita_mit | 0 = no measure<br>1 + n = cumulative number of relaxed measures | Cumulative number of relaxed kindergarten and nursery school measures over time. The first relaxation is given the value 1. All subsequent new relaxations become 1 + n number of previous relaxations. |
|  |  | kita_res_mit | 1 = zero point | Interval scaled variable measuring the change in present restricted and relaxed measures. If there is a restriction, the value is increased by 0.01 points or decreased by this value if there is a relaxation. |
|  |  | kita_closure | 0 = no measure in place<br>1 = closure<br>2 = opening | Categorical measure of kindergarten and nursery school closures. |
| Public institutions | **University/**<br>**higher education** | university | 0 = no measure<br>1 = restricted measure<br>2 = relaxed measure<br>3 = both measures in place | General restricted and relaxed measures in universities and facilities of higher education.<br>"3 = both measures in place" means that both restricted and relaxed measures are present at the same time. |
|  |  | university_res | 0 = no measure<br>1 + n = cumulative number of restricted measures | Cumulative number of restricted universities and facilities of higher education measures over time. The first restriction is given the value 1. All subsequent new restrictions become 1 + n number of previous restrictions. |
|  |  | university_mit | 0 = no measure<br>1 + n = cumulative number of relaxed measure | Cumulative number of relaxed universities and facilities of higher education measures over time. The first relaxation is given the value 1. All subsequent new relaxations become 1 + n number of previous relaxations. |
|  |  | university_res_mit | 1 = zero point | Interval scaled variable measuring the change in present restricted and relaxed measures. If there is a restriction, the value is increased by 0.01 points or decreased by this value if there is a relaxation. |
|  |  | university_closure | 0 = no measure in place<br>1 = closure<br>2 = opening | Categorical measure of universities and facilities of higher education closures. |
| Public institutions & Public spaces | **Cultural and educational institutions** | culture | 0 = no measure<br>1 = restricted measure<br>2 = relaxed measure<br>3 = both measures in place | General restricted and relaxed measures in cultural and educational institutions.<br>"3 = both measures in place" means that both restricted and relaxed measures are present at the same time. |
|  |  | culture_res | 0 = no measure<br>1 + n = cumulative number of restricted measures | Cumulative number of restricted cultural and educational institutions measures over time. The first restriction is given the value 1. All subsequent new restrictions become 1 + n number of previous restrictions. |
|  |  | culture_mit | 0 = no measure<br>1 + n = cumulative number of relaxed measures | Cumulative number of relaxed cultural and educational institutions measures over time. The first relaxation is given the value 1. All subsequent new relaxations become 1 + n number of previous relaxations. |
|  |  | culture_res_mit | 1 = zero point | Interval scaled variable measuring the change in present restricted and relaxed measures. If there is a restriction, the value is increased by 0.01 points or decreased by this value if there is a relaxation. |
|  |  | culture_closure | 0 = no measure in place<br>1 = closure<br>2 = opening | Categorical measure of cultural and educational institutions closures. |
| Public institutions | **Workplace restrictions** | workplace | 0 = no measure<br>1 = restricted measure<br>2 = relaxed measure<br>3 = both measures in place | General restricted and relaxed measures in workplace.<br>"3 = both measures in place" means that both restricted and relaxed measures are present at the same time. |
|  |  | workplace_res | 0 = no measure<br>1 + n = cumulative number of restricted measures | Cumulative number of restricted workplace measures over time. The first restriction is given the value 1. All subsequent new restrictions become 1 + n number of previous restrictions. |
|  |  | workplace_mit | 0 = no measure<br>1 + n = cumulative number of relaxed measures | Cumulative number of relaxed workplace measures over time. The first relaxation is given the value 1. All subsequent new relaxations become 1 + n number of previous relaxations. |
|  |  | workplace_res_mit | 1 = zero point | Interval scaled variable measuring the change in present restricted and relaxed measures. If there is a restriction, the value is increased by 0.01 points or decreased by this value if there is a relaxation. |

## References

1 WHO | Non-pharmaceutical public health measures for mitigating the risk and impact of epidemic and pandemic influenza. World Health Organization, 2019 http://www.who.int/influenza/publications/public_health_measures/publication/en/ (accessed Nov 6, 2020).

2 Porcher S. Response2covid19, a dataset of governments’ responses to COVID-19 all around the world. Sci Data 2020; 7: 423.

3 Cheng C, Barceló J, Hartnett AS, Kubinec R, Messerschmidt L. COVID-19 Government Response Event Dataset (CoronaNet v.1.0). Nat Hum Behav 2020; 4: 756–68.

4 Zheng Q, Jones FK, Leavitt S V, et al. HIT-COVID, a global database tracking public health interventions to COVID-19. Sci Data 2020; 7: 286.

5 Desvars-Larrive A, Dervic E, Haug N, et al. A structured open dataset of government interventions in response to COVID-19. medRxiv 2020;: 2020.05.04.20090498.

6 The European Observatory on Health Systems and Policies. www.covid19healthsystem.org.

7 Hale T, Webster S, Petherick A, Phillips T, Kira B. Oxford COVID-19 Government Response Tracker, Blavatnik School of Government. 2020.

